# Longitudinal Analysis Reveals Distinct Antibody and Memory B Cell Responses in SARS-CoV2 Naïve and Recovered Individuals Following mRNA Vaccination

**DOI:** 10.1101/2021.03.03.21252872

**Authors:** Rishi R. Goel, Sokratis A. Apostolidis, Mark M. Painter, Divij Mathew, Ajinkya Pattekar, Oliva Kuthuru, Sigrid Gouma, Leticia Kuri-Cervantes, Wenzhao Meng, Sharon Adamski, Amy E. Baxter, Josephine R. Giles, Madison E. Weirick, Christopher M. McAllister, Amanda Hicks, Scott Korte, Jeanette Dougherty, Sherea Long, Kurt D’Andrea, Jacob T. Hamilton, Eline T Luning Prak, Michael R. Betts, Paul Bates, Scott E. Hensley, Allison R. Greenplate, E. John Wherry

## Abstract

Novel mRNA vaccines for SARS-CoV2 have been authorized for emergency use and are currently being administered to millions of individuals worldwide. Despite their efficacy in clinical trials, there is limited data on vaccine-induced immune responses in individuals with a prior SARS-CoV2 infection compared to SARS-CoV2 naïve subjects. Moreover, how mRNA vaccines impact the development of antibodies as well as memory B cells in COVID-19 experienced versus COVID-19 naïve subjects remains poorly understood. In this study, we evaluated antibody responses and antigen-specific memory B cell responses over time in 33 SARS-CoV2 naïve and 11 SARS-CoV2 recovered subjects. mRNA vaccination induced significant antibody and memory B cell responses against full-length SARS-CoV2 spike protein and the spike receptor binding domain (RBD). SARS-CoV2 naïve individuals benefitted from both doses of mRNA vaccine with additional increases in antibodies and memory B cells following booster immunization. In contrast, SARS-CoV2 recovered individuals had a significant immune response after the first dose with no increase in circulating antibodies or antigen-specific memory B cells after the second dose. Moreover, the magnitude of the memory B cell response induced by vaccination was lower in older individuals, revealing an age-dependence to mRNA vaccine-induced B cell memory. Side effects also tended to associate with post-boost antibody levels, but not with post-boost memory B cells, suggesting that side effect severity may be a surrogate of short-term antibody responses. The frequency of pre-vaccine antigen-specific memory B cells in SARS-CoV2 recovered individuals strongly correlated with post-vaccine antibody levels, supporting a key role for memory B cells in humoral recall responses to SARS-CoV2. This observation may have relevance for future booster vaccines and for responses to viral variants that partially escape pre-existing antibodies and require new humoral responses to be generated from memory B cells. Finally, post-boost antibody levels were not correlated with post-boost memory responses in SARS-CoV2 naïve individuals, indicating that short-term antibody levels and memory B cells are complementary immunological endpoints that should be examined in tandem when evaluating vaccine response. Together, our data provide evidence of both serological response and immunological memory following mRNA vaccination that is distinct based on prior SARS-CoV2 exposure. These findings may inform vaccine distribution in a resource-limited setting.

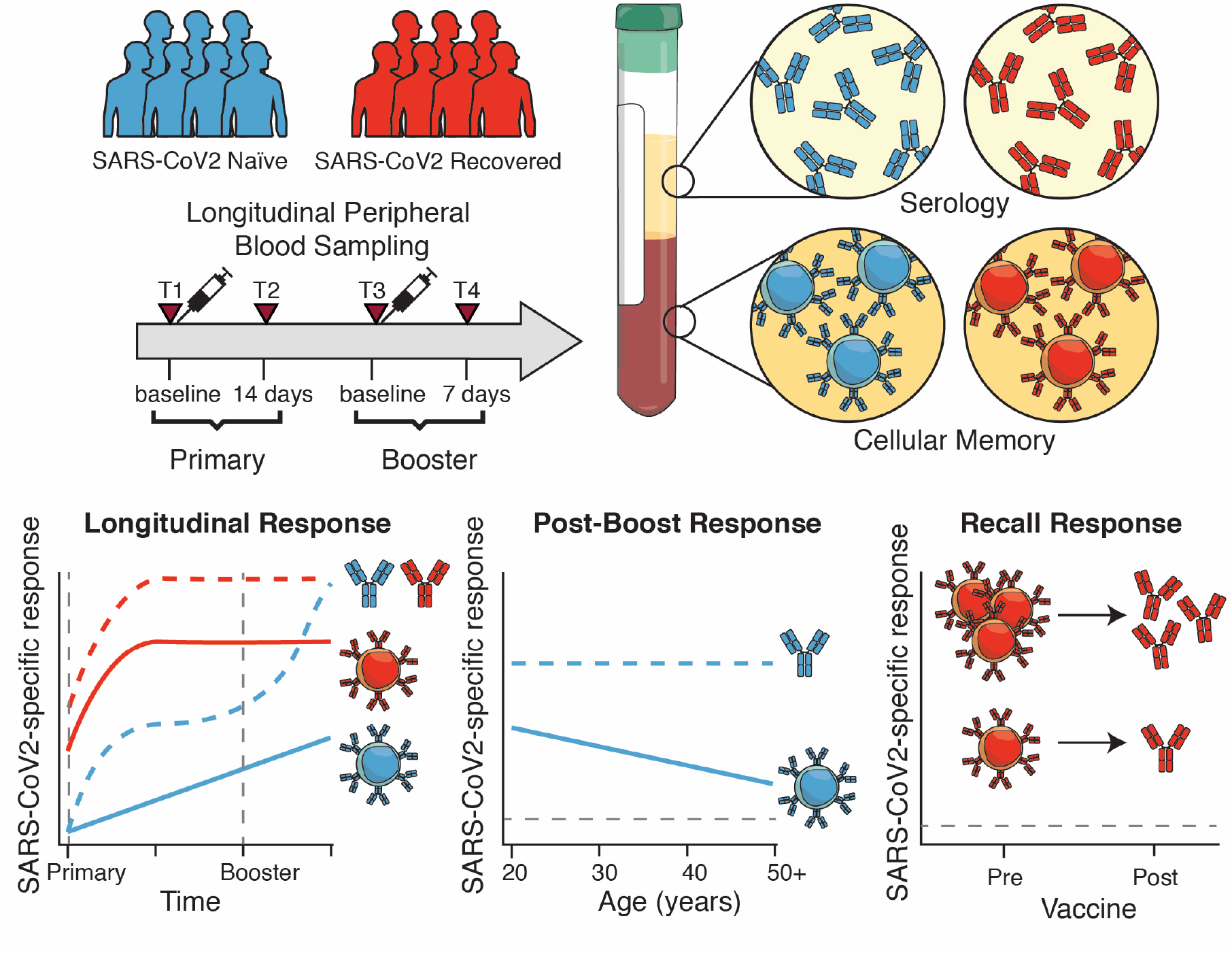

## INTRODUCTION

The COVID-19 pandemic has resulted in more than 100 million infections and 2.5 million deaths worldwide. Novel vaccines have recently been issued emergency use authorization by the FDA and are currently being administered to front-line workers and at-risk individuals. Early data from clinical trials suggest that these vaccines are safe and effective^1,2^, however there is still a paucity of information interrogating how these novel mRNA vaccines elicit immune responses at the cellular and molecular level.

The humoral immune response to infection or vaccination results in two major outcomes: the production of antibodies by antibody secreting cells (ASC), which can provide rapid protective immunity, and the generation of long-lived memory B cells capable of mounting recall responses^3^. If circulating antibodies fail to confer protection to a future exposure, memory B cells drive the recall response by producing new antibodies through formation of new ASC or re-initiating germinal center reactions to generate new high-affinity B cell clones through additional rounds of somatic hypermutation. In the context of acute SARS-CoV2 infection, immunological memory in the form of antibodies and memory B cells has been shown to be durable for over 8 months post-symptom onset^4–6^. However, studies on vaccinated individuals have largely focused on measuring binding and/or neutralizing antibodies as primary endpoints^7,8^. Although antibodies are a central component of vaccine efficacy, immunological memory in the form of memory B cells may be important for long-term protection, responses to subsequent infection, and the ability to respond to emerging variant strains. The induction of memory B cells by mRNA vaccines remains poorly understood. Furthermore, it is unclear how memory B cell responses relate to serological responses, and how both antibody and memory B cell responses differ in subjects who previously experienced SARS-CoV2 infection versus those who are SARS-CoV2 naïve.

A related question is whether individuals who experienced prior SARS-CoV2 infection require a second dose of mRNA vaccine. This question is particularly important given the currently limited vaccine supply and challenges with deployment. As these individuals have already generated a primary immune response to SARS-CoV2 during their natural infection, it is possible that a single dose of vaccine could be enough to sufficiently boost their antibody and memory B cell responses. Indeed, several recent studies have indicated that antibody responses can be robustly induced in SARS-CoV2 experienced individuals, consistent with an anamnestic response^9–12^. Although one study suggests that memory B cells might also be boosted after a single vaccine dose^13^, it remains unclear how well memory B cell responses are induced in SARS-CoV2 naïve versus SARS-CoV2 experienced subjects after one versus two doses of mRNA vaccine. Moreover, how antibody levels predict or relate to memory B cell responses following mRNA vaccination remains to be determined. These key gaps in our understanding require longitudinal analysis of antibodies together with memory B cell responses after the first and second dose of mRNA vaccine in SARS-CoV2 naïve and experienced subjects.

In this study, we established a longitudinal cohort of SARS-CoV2 naïve and SARS-CoV2 recovered individuals who received mRNA vaccines at the University of Pennsylvania Health System. From these longitudinal samples, we assessed both circulating antibodies and antigen-specific memory B cells over the course of first and second immunization. We further integrated these serologic and cellular assays with clinical metadata and compared these immune responses with those from non-vaccinated SARS-CoV2 recovered subjects. These studies revealed several key findings. First, as others have reported, vaccination boosts antibody levels more quickly in SARS-CoV2 recovered versus naïve subjects. Second, memory B cell responses are also robustly induced by the first dose of vaccine in SARS-CoV2 recovered subjects, but no additional boosting is observed after the second vaccine dose. In contrast, memory B cell responses continue to improve after the second vaccination in SARS-CoV2 naive subjects. Third, although subjects of all ages benefit from induction of serological and cellular immunity, vaccine induction of memory B cells declines with age. Fourth, there was a trend for mRNA vaccine-induced antibody levels to be higher in subjects with more systemic side effects, but side effects had no relation to memory B cell responses. Finally, there was no relationship between post-vaccination serum antibody and memory B cells in SARS-CoV2 naïve subjects, indicating that measuring short-term antibody titers alone may fail to predict long-term immunity elicited by the vaccine. Pre-existing memory B cells did strongly correlate with post-vaccination antibody responses in SARS-CoV2 recovered subjects, further emphasizing the importance of measuring these cells. These data highlight the efficacy of SARS-CoV2 mRNA vaccines and support a single-dose vaccine regimen in SARS-CoV2 recovered subjects, which may allow more effective distribution of vaccines to the general population.

## METHODS

### Recruitment and Clinical Sample Collection

44 individuals (33 SARS-CoV2 naïve, 11 SARS-CoV2 recovered) were consented and enrolled in the study with approval from the University of Pennsylvania Institutional Review Board (IRB# 844642). All subjects received either Pfizer (BNT162b2) or Moderna (mRNA-1273) mRNA vaccines. Samples were collected at 4 timepoints: baseline, 2 weeks post-primary immunization, day of booster immunization, and 1 week post-booster immunization. 80-100mL of peripheral blood samples and clinical questionnaire data were collected at each study visit. Full cohort and demographic information is provided in **figure S1**. Non-vaccinated recovered COVID-19 donors (RD) were adults with a prior positive COVID-19 PCR test by self-report who met the definition of recovery by the Centers for Disease Control^14^.

### Sample Processing

Venous blood was collected into sodium heparin and EDTA tubes by standard phlebotomy. Blood tubes were centrifuged at 3000rpm for 15 minutes to separate plasma. Heparin and EDTA plasma were stored at −80C for downstream antibody analysis. Remaining whole blood was diluted 1:1 with RPMI + 1% FBS and layered onto SEPMATE tubes (STEMCELL Technologies) containing lymphoprep gradient (STEMCELL Technologies). SEPMATE tubes were centrifuged at 1200g for 10 minutes and the PBMC fraction was collected into new tubes. PBMCs were then washed with RPMI + 1% FBS and treated with ACK lysis buffer (Thermo Fisher) for 5 minutes. Samples were washed again with RPMI + 1% FBS, filtered with a 70um filter, and counted using a Countess automated cell counter (Thermo Fisher). Aliquots containing 5×10^6^ PBMCs were cryopreserved in 90% FBS 10% DMSO.

### Detection of SARS-CoV2-Specific Antibodies

Plasma samples were tested for SARS-CoV2-specific antibody by enzyme-linked immunosorbent assay (ELISA) as previously described^15^. Plasmids encoding the recombinant full-length spike protein and the receptor binding domain (RBD) were provided by F. Krammer (Mt. Sinai) and purified by nickel-nitrilotriacetic acid resin (Qiagen). ELISA plates (Immulon 4 HBX, Thermo Fisher Scientific) were coated with PBS or 2 ug/mL recombinant protein and stored overnight at 4C. The next day, plates were washed with phosphate-buffered saline containing 0.1% Tween-20 (PBS-T) and blocked for 1 hour with PBS-T supplemented with 3% non-fat milk powder. Samples were heat-inactivated for 1 hour at 56C and diluted in PBS-T supplemented with 1% non-fat milk powder. After washing the plates with PBS-T, 50 uL diluted sample was added to each well. Plates were incubated for 2 hours and washed with PBS-T. Next, 50 uL of 1:5000 diluted goat anti-human IgG-HRP (Jackson ImmunoResearch Laboratories) or 1:1000 diluted goat anti-human IgM-HRP (SouthernBiotech) was added to each well and plates were incubated for 1 hour. Plates were washed with PBS-T before 50 uL SureBlue 3,3’,5,5’-tetramethylbenzidine substrate (KPL) was added to each well. After 5 minutes incubation, 25 uL of 250 mM hydrochloric acid was added to each well to stop the reaction. Plates were read with the SpectraMax 190 microplate reader (Molecular Devices) at an optical density (OD) of 450 nm. Monoclonal antibody CR3022 was included on each plate to convert OD values into relative antibody concentrations. Plasmids to express CR3022 were provided by I. Wilson (Scripps).

### Detection of SARS-CoV2-Specific Memory B Cells

Antigen-specific B cells were detected using biotinylated proteins in combination with different streptavidin (SA)-fluorophore conjugates^4,6^. Biotinylated proteins were multimerized with fluorescently labeled SA for 1 hour at 4C. Full-length spike protein (R&D Systems) was mixed with SA-BV421 (Biolegend) at a 10:1 mass ratio (e.g., 200ng spike with 20ng SA; ∼4:1 molar ratio). Spike RBD (R&D Systems) was mixed with SA-APC (Biolegend) at a 2:1 mass ratio (e.g., 25ng RBD with 12.5ng SA; ∼4:1 molar ratio). Biotinylated influenza HA pools were mixed with SA-PE (Biolegend) at a 6.25:1 mass ratio (e.g., 100ng HA pool with 16ng SA; ∼6:1 molar ratio). Individual influenza HA antigens corresponding with the 2019 trivalent vaccine (A/Brisbane/02/2018/H1N1, B/Colorado/06/2017; Immune Technology) were biotinylated using an EZ-Link Micro NHS-PEG4 Biotinylation Kit (Thermo Fisher) according to the manufacturer’s instructions. Excess biotin was subsequently removed using Zebra Spin Desalting Columns 7K MWCO (Thermo Fisher) and protein was quantified with a Pierce BCA Assay (Thermo Fisher). SA-BV711 (BD Bioscience) was used as a decoy probe without biotinylated protein to gate out cells that non-specifically bind streptavidin. All experimental steps were performed in a 50/50 mixture of PBS + 2% FBS and Brilliant Buffer (BD Bioscience). Antigen probes for spike, RBD, and HA were prepared individually and mixed together after multimerization with 5uM free D-biotin (Avidity LLC) to minimize potential cross-reactivity between probes. For staining, 5×10^6^ cryopreserved PBMC samples were prepared in a 96-well U-bottom plate. Cells were first stained with Fc block (Biolegend, 1:200) and Ghost 510 Viability Dye (Tonbo Biosciences, 1:600) for 15 minutes at 4C. Cells were then washed and stained with 50uL antigen probe master mix containing 200ng spike-BV421, 25ng RBD-APC, 100ng HA-PE, and 20ng SA-BV711 decoy for 1 hour at 4C. Following incubation with antigen probe, cells were washed again and stained with anti-CD3 (BD Bioscience, 1:200), anti-CD19 (Biolegend, 1:100), anti-CD20 (BD Bioscience, 1:500), anti-CD27 (BD Bioscience, 1:200), anti-CD38 (BD Bioscience, 1:200), anti-CD71 (BD Bioscience, 1:50), anti-IgD (BD Bioscience, 1:50), anti-IgM (Biolegend, 1:200), and anti-IgG (Biolegend, 1:400). After surface stain, cells were washed and fixed in 1% PFA overnight at 4C.

### Flow Cytometry

Samples were acquired on a BD Symphony A5 instrument. Standardized SPHERO rainbow beads (Spherotech) were used to track and adjust photomultiplier tubes over time. UltraComp eBeads (Thermo Fisher) were used for compensation. Up to 5×10^6^ cells were acquired per sample. Data were analyzed using FlowJo v10 (BD Bioscience). Full gating strategy is shown in **figure S2**.

### Data Visualization and Statistics

All data were analyzed using custom R scripts. Statistical tests are indicated in the corresponding figure legends. *indicates p < 0.05, **indicates p < 0.01, ***indicates p < 0.001, ****indicates p < 0.0001. Source code and data files are available upon request from the authors.

## RESULTS

For this study, we recruited 44 individuals who received SARS-CoV2 mRNA vaccines (Pfizer BNT162b2 or Moderna mRNA-1273) at the University of Pennsylvania Health System. Of this cohort, 11 individuals had a prior SARS-CoV2 infection. Peripheral blood samples were collected for immunological analysis at 4 key timepoints (**figure 1A**): pre-vaccine baseline (timepoint 1), 2 weeks following the first dose (timepoint 2), the day of second dose (timepoint 3), and 1 week following the second dose (timepoint 4). This study design allowed us to investigate the kinetics of immune responses following both primary and secondary immunizations.

**Figure 1.**
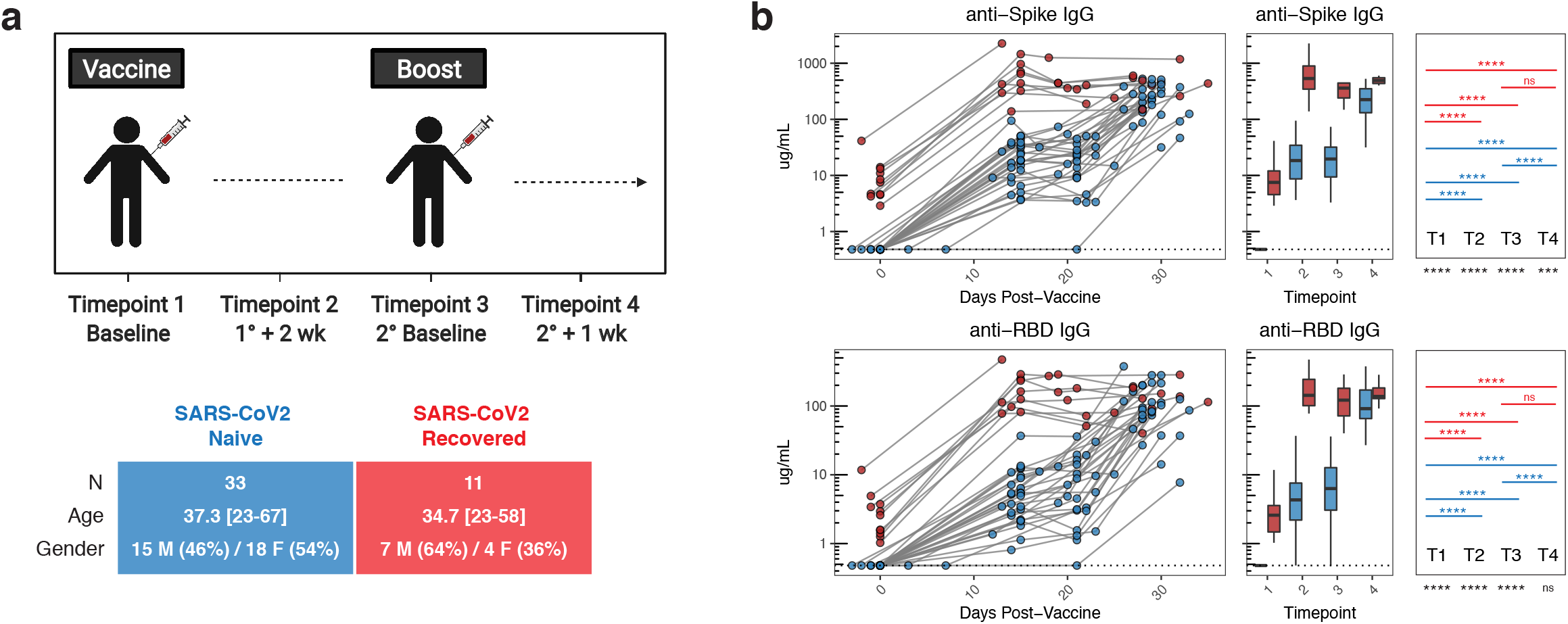
Antibody responses following mRNA vaccination in SARS-CoV2 naïve and SARS-CoV2 recovered individuals. **A)** UPenn Immune Health COVID vaccine study design. **B)** Concentration of anti-spike and anti-RBD IgG antibodies in vaccinated individuals over time. Dotted lines indicate the limit of detection for the assay. Blue = SARS-CoV2 naïve + mRNA vaccine, red = SARS-CoV2 recovered + mRNA vaccine. Statistics were calculated using unpaired Kruskal-Wallis or Wilcoxon test with adjustment for multiple comparisons.

We first measured circulating antibody responses in longitudinal serum samples by ELISA. At baseline, SARS-CoV2 naïve individuals had undetectable levels of IgG antibodies specific for either full-length spike protein or the spike receptor binding domain (RBD) (**figure 1B**). Primary vaccination induced a significant increase in SARS-CoV2-specific antibodies, that was further enhanced by the booster dose (**figure 1B**). In contrast, all SARS-CoV2 recovered individuals had detectable levels of anti-spike and anti-RBD IgG at baseline and these antibody responses were significantly increased after the first dose of vaccine (**figure 1B**). However, in SARS-CoV2 recovered subjects, there was no additional increase in antibody levels following the second vaccine dose (**figure 1B**). Notably, the levels of anti-RBD IgG were similar in the SARS-CoV2 naïve and SARS-CoV2 recovered individuals at 1 week post-boost (timepoint 4) (**figure 1B**).

We next asked how mRNA vaccination impacted the responses of memory B cells specific for SARS-CoV2. To address this question, we developed a flow cytometric assay using fluorescently labeled antigens as probes to track induction of virus-specific memory B cells in longitudinal PBMC samples (**figure 2A**). Consistent with the antibody data, SARS-CoV2 naïve individuals had minimal spike-specific memory B cells at baseline, whereas SARS-CoV2 recovered individuals had a significant population of spike-specific memory B cells ranging from ∼0.15-0.8% of total memory B cells (**figure 2B**). The frequency of these antigen-specific memory B cells was comparable to a separate cohort of non-vaccinated SARS-CoV2 recovered donors (**figure 2B**). Similar trends were observed for memory B cells targeting the spike RBD (**figure 2B**). After primary immunization, SARS-CoV2 naïve individuals had a significant increase in spike-specific and RBD-specific memory B cells over baseline (**figure 2B**). These memory B cells were also significantly boosted after adminstration of the second dose, approaching the levels of memory B cells observed in non-vaccinated SARS-CoV2 recovered donors (**figure 2B**). In contrast, SARS-CoV2 recovered individuals had a robust expansion of spike- and RBD-specific memory B cells following primary immunization, but had no additional boosting after the second vaccine dose (**figure 2B**), suggesing minimal benefit of the second dose in these recovered subjects. As a control we also examined the frequency of influenza hemagglutinin (HA)-specific memory B cells in both SARS-CoV2 naïve and recovered individuals following SARS-CoV2 vaccination. The frequency of these antigen-unrelated memory B cells remained stable throughout the mRNA vaccination timecourse (**figure 2B**), confirming the specificity of this memory B cell assay. Together, these results demonstrate robust induction of SARS-CoV2-specific memory B cells by two doses of mRNA vaccine in SARS-CoV2 naïve subjects. Alternatively, a single dose of mRNA vaccine amplified pre-existing antigen-specific memory B cells in SARS-CoV2 recovered subjects, with no additional quantitative benefit after the second vaccine dose.

**Figure 2.**
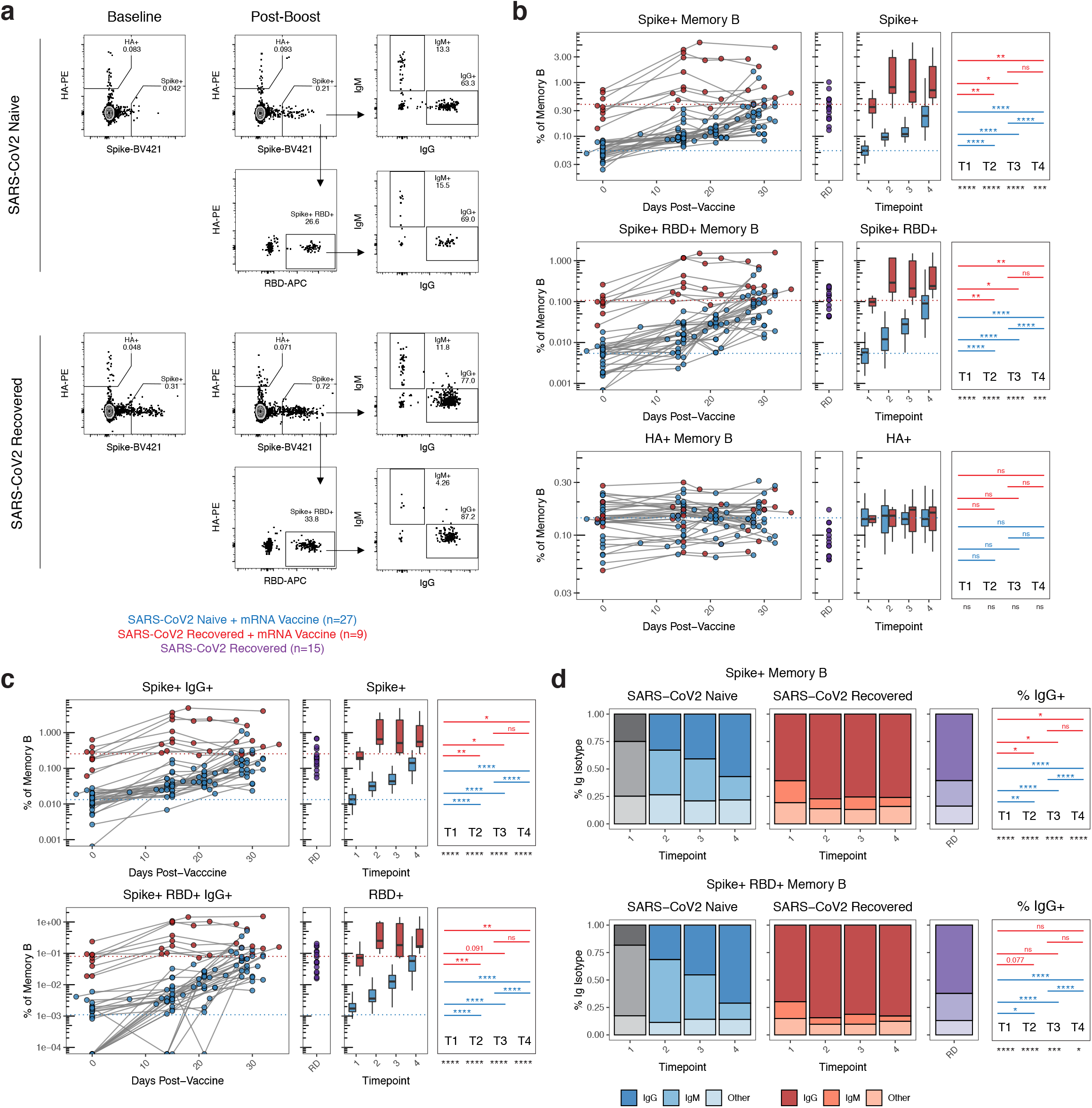
Antigen-specific memory B cell responses following mRNA vaccination in SARS-CoV2 naïve and SARS-CoV2 recovered individuals. **A)** Gating strategy and representative plots for flow cytometric analysis of SARS-CoV2-specific B cells. Cells were stained with fluorescently labeled SARS-CoV2 full-length spike protein, SARS-CoV2 spike receptor binding domain (RBD), and influenza hemagglutinin (HA). Memory B cells were identified as live, CD3^-^, CD19^+^, non-naïve (≠ IgD^+^ CD27^-^), CD20^+^ CD38^lo/int^, decoy^-^ cells. Spike^+^ HA^-^ cells were subsequently analyzed for binding to RBD, as well as immunoglobulin class (IgG vs. IgM). **B)** Frequency of spike^+^, spike^+^/RBD^+^, and HA^+^ memory B cells over time in vaccinated individuals. Data are represented as frequency of antigen-specific cells in the total memory B cell compartment. **C)** Frequency of antigen-specific IgG^+^ memory B cells over time in vaccinated individuals. Data are represented as frequency of antigen-specific IgG^+^ cells in the total memory B cell compartment. **D)** Frequency of IgG and IgM isotypes over time in the antigen-specific memory cell compartments. RD = non-vaccinated, SARS-CoV2 recovered donors. Dotted lines indicate the mean at baseline in SARS-CoV2 naïve and SARS-CoV2 recovered individuals. Blue = SARS-CoV2 naïve + mRNA vaccine, red = SARS-CoV2 recovered + mRNA vaccine, purple = SARS-CoV2 recovered. Statistics were calculated using unpaired Kruskal-Wallis or Wilcoxon test with adjustment for multiple comparisons.

We further analyzed the immunoglobulin isotype of SARS-CoV2 specific memory B cells. On day 15 after primary immunization, ∼25-30% of responding spike-specific memory B cells were IgG^+^ and ∼40-50% were IgM^+^ in SARS-CoV2 naïve individuals (**figure 2C**). The frequency of IgG^+^ memory B cells increased to >50% following the second dose of vaccine in these subjects (**figure 2C-D**), consistent with a qualitative improvement in B cell memory formation after the boost. Conversely, in SARS-CoV2 recovered individuals, ∼60-70% of spike-specific memory B cells detected prior to vaccination were IgG^+^ (**figure 2C-D**). Although the frequency of IgG^+^ memory B cells increased slightly to ∼75% following the first dose of vaccine, further increases were not observed after the second immunization (**figure 2C-D**). A similar pattern of IgG frequency was observed for RBD-specific memory B cells (**figure 2C-D**). In addition, the fraction of spike-specific memory B cells that recognized RBD remained stable over time in SARS-CoV2 recovered individuals. In SARS-CoV2 naïve subjects, the fraction of the overall spike-specific memory B cell response that was focused on RBD increased over time, becoming equivalent to that observed in SARS-CoV2 recovered individuals after the second vaccine dose (**figure S2**). Overall, these data indicate a qualitative benefit to the virus-specific memory B cell response following both doses of vaccine in SARS-CoV2 naïve individuals, and limited qualitative improvement following the first but not the second vaccine dose in SARS-CoV2 recovered subjects.

Several previous studies have reported a negative association between age and vaccine-induced antibody titers after a single dose of mRNA vaccines^16,17^. We therefore investigated potential relationships between sex or age and B cell responses after one or two doses of vaccine. In our cohort of SARS-CoV2 naïve vaccinees, there were no associations between sex and either antibody or memory B cell responses (**figure 3A, 3C**). Although there was no association between age and spike-specific IgG after the first immunization (i.e. pre-boost), there was a modest trend towards a negative relationship between RBD-specific IgG titers and age after the first vaccine dose (**figure 3B**). There was no significant correlation between age and either spike- or RBD-specific serum IgG after the second dose (**figure 3B**). In contrast, there was a clear negative correlation between the post-boost frequency of antigen-specific memory B cells and age (**figure 3D**). Although this relationship represented weaker induction of memory B cells with older age, all age groups still displayed an increase in the frequency of SARS-CoV2 specific memory B cells compared to pre-vaccine baseline (**figure S3A-D**). There was also no change in the frequency of total memory B cells by sex or age, indicating the antigen-specific nature of this effect (**figure S3E**). While our cohort is not extensively enriched in those over 50 years old, and does not directly address elderly vaccinees, age associations with weaker vaccine-induced antibody responses appeared to normalize following the second dose of vaccine. Conversely, the effect of age on memory B cell responses was more prominent after the second immunization. These data point to potentially relevant age-related changes in immune response to vaccination.

**Figure 3.**
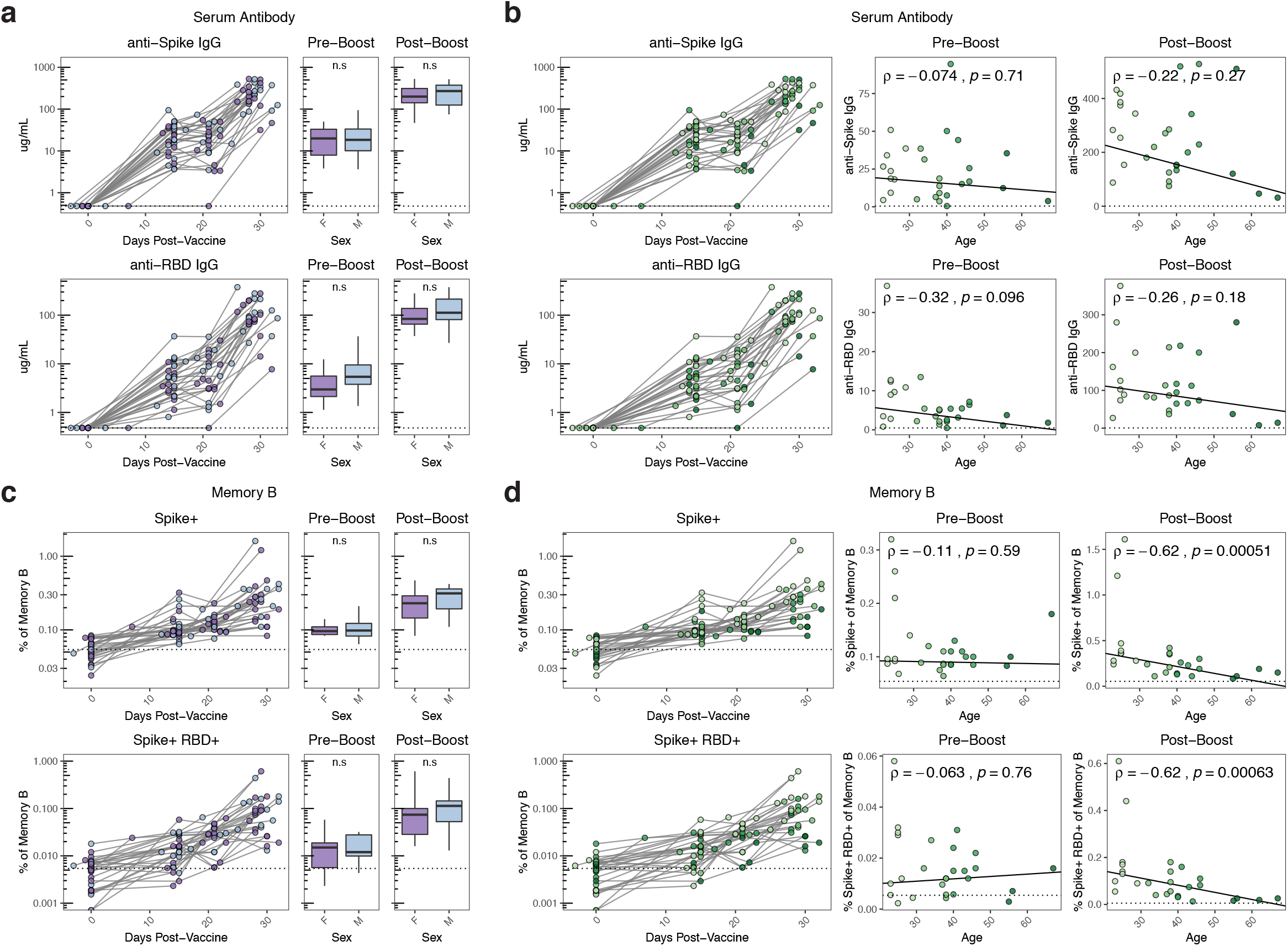
Age-associated decreases in antigen-specific B cell responses following mRNA vaccination. **A, B)** Concentration of anti-spike and anti-RBD IgG antibodies over time compared with sex and age in SARS-CoV2 naïve individuals. Dotted lines indicate the limit of detection for the assay. **C, D)** Frequency of spike^+^ and spike^+^/RBD^+^ memory B cells over time compared with sex and age in SARS-CoV2 naïve individuals. Data are represented as frequency of spike^+^ and spike^+^/RBD^+^ cells in the total memory B cell compartment. Pre-boost indicates samples collected at timepoint 2 (∼15 days post-primary vaccination). Post-boost indicates samples collected at timepoint 4 (∼7 days post-secondary vaccination). Dotted lines indicate the mean frequency of cells at baseline. Statistics for sex were calculated using Wilcoxon test. Associations with age were calculated using Spearman correlation.

An additional question is whether vaccine-induced side effects have any relationship to immune responses^9^. We addressed this question by comparing vaccine-induced antibody and memory B cell responses in subjects with or without self-reported systemic side effects (i.e. fever, chills, headache, fatigue, myalgia). In SARS-CoV2 naïve vaccinees with systemic side-effects following the second dose, there was a trend towards increase in antibody responses at the post-boost timepoint (**figure S4**). Such a trend was not observed for the memory B cell response. Although these data only represent a statistical trend, they do provoke questions about potential relationships between early vaccine-induced inflammation and the induction of antibody responses, which should be addressed in future studies.

Finally, we investigated the potential relationships between antibody and memory B cell responses. In SARS-CoV2 naïve subjects, we examined the relationship between circulating antibody responses and memory B responses after two doses of vaccine. Despite strong induction of both spike- and RBD-specific antibody and memory B cells in these subjects, there was no association between the levels of post-boost antibodies and B cell memory (**figure 4A**), indicating that short-term serologies and cellular memory are distinct immunological measures of vaccine efficacy. Similarly, pre-vaccine baseline antibody levels did not correlate with baseline memory B cell frequencies in SARS-CoV2 recovered individuals (**figure 4B**). We next asked which measure of humoral immunity predicted antibody recall responses post-vaccination. Interestingly, the baseline levels of SARS-CoV2-specific antibody did not correlate with the level of antibody achieved after primary vaccine (timepoint 2) in SARS-CoV2 recovered donors (**figure 4C**). However, the baseline frequency of antigen-specific memory B cells (timepoint 1) strongly correlated with post-primary vaccine antibody levels (timepoint 2, **figure 4D)**, consistent with the notion that these pre-vaccination memory B cells are major contributors to the SARS-CoV2 antibody recall response. Overall, these data highlight the importance of measuring antigen-specific memory B cells in addition to more conventional serologic approaches as an immunological correlate of vaccine-induced immunity.

**Figure 4.**
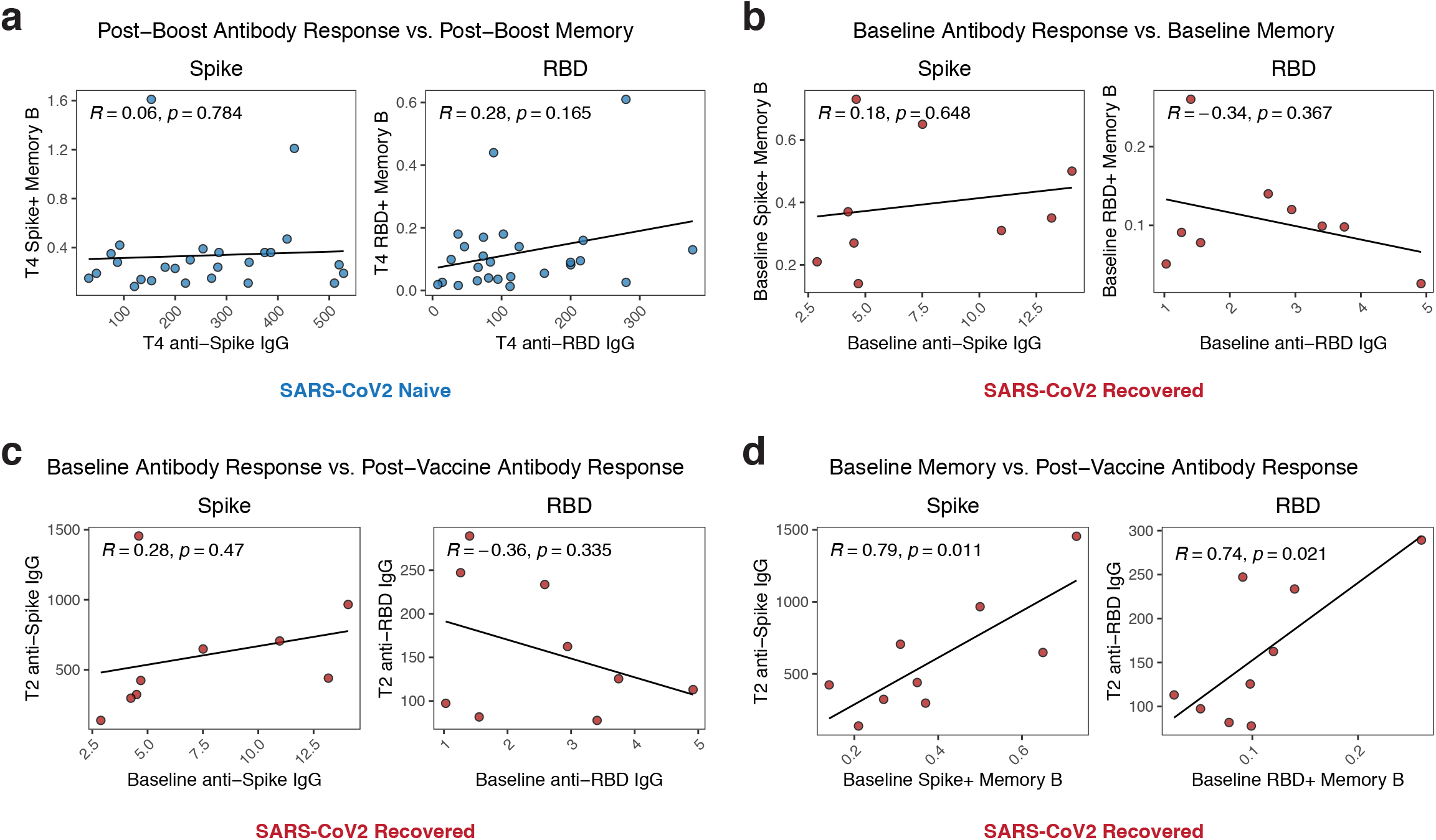
Antigen-specific memory cells are a distinct measure of vaccine efficacy and correlate with antibody recall responses. **A)** Association of post-boost (timepoint 4) antibody levels with post-boost (timepoint 4) antigen-specific memory cell frequencies in SARS-CoV2 naïve individuals. **B)** Association of baseline (timepoint 1) antibody levels with baseline (timepoint 1) antigen-specific memory cell frequencies in SARS-CoV2 recovered individuals. **C)** Association of baseline (timepoint 1) antibody levels with post-primary vaccination (timepoint 2) antibody levels in SARS-CoV2 recovered individuals. **D)** Association of baseline (timepoint 1) antigen-specific memory cell frequencies with post-primary vaccination (timepoint 2) antibody levels in SARS-CoV2 recovered individuals. Associations between immunological parameters were calculated using Pearson correlation.

## DISCUSSION

Our data indicate that mRNA vaccines induce significant antibody and memory B cell responses to full-length spike and the RBD. These results are encouraging for both short- and long-term vaccine efficacy. Overall, these data also add to our understanding of SARS-CoV2 mRNA vaccine-induced immune responses in several ways. First, our serological data is consistent with several other recent studies^9,10,12,13,16,17^ indicating robust boosting of antibody responses in SARS-CoV2 recovered subjects after the first vaccine dose, but little benefit to antibody titers after the second vaccine dose. Moreover, we identified a similar effect for virus-specific memory B cells, demonstrating that both a quantitative and qualitative plateau in vaccine-induced memory B cells is achieved following the first dose of vaccine with little additional change to the memory B cell response following booster vaccination. These data advocate for only a single vaccine dose in individuals confirmed to have previously been infected with SARS-CoV2. It is important, however, to point out that our cohort consisted of individuals who were not hospitalized during their SARS-CoV2 infections, and it may be necessary to address this question in individuals who experienced more severe COVID-19.

It remains unclear if the second vacccine dose in recovered individuals has other immunological effects not reflected in overall antibody titers or memory B cell frequencies, such as expansion of specific, high-affinity B cell clones. Additional analysis of BCR sequences and memory B cell differentiation states is necessary to fully address these questions. It is also possible that booster vaccination has some beneficial effects on virus-specific T cell responses in SARS-CoV2 recovered individuals, and this topic should also be investigated. A second related point is that in SARS-CoV2 naïve individuals, both the antibody response and the memory B cell response displayed considerable benefit from the second dose of mRNA vaccine. It is possible that some of this serological and memory B cell maturation would occur over time in the absence of a booster vaccination, but the spike- and RBD-specific antibody titers appeared to plateau between the first and second doses of vaccine. Moreover, the frequency of the memory B cell response that was IgG^+^ and the fraction of the overall spike-specific memory B cell response that was focused on RBD both improved after booster vaccination, arguing strongly for the benefit of a two dose mRNA vaccine schedule in SARS-CoV2 naïve individuals.

In this cohort we also observed a negative association of age with induction of B cell memory. Although others have reported a negative association between age and serum antibody titers after a single mRNA vaccine dose^16,17^, we found that this relationship was not significant following two doses of mRNA vaccination. However, we observed that the magnitude of the memory B cell response following the second dose was lower with increased age, confirming age as a key variable in mRNA vaccine induced immunity. It remains unclear if the age-associated effect on memory B cell induction represents a true difference in the magnitude of response or a difference in kinetics that will resolve at later timepoints. It is also challenging to define an exact threshold for how much immunological memory is sufficient to provide long-term protection. Although all subjects, regardless of age, had significant humoral and memory B cell responses to vaccination, these data highlight a need to further understand the age-related changes in responses to mRNA vaccination. In examining correlates of vaccine-induced immune responses, we also uncovered a trend suggesting that vaccine-induced side effects may be related to post-vaccination serum antibodies, but not memory B cells. While more data are needed, it is possible that systemic inflammation early after vaccination could contribute to an initial induction of antibody with less of an impact on the development of memory B cells. Larger cohorts and more quantitative measures of vaccine-induced side effects may further clarify these relationships.

Finally, our data demonstrate the importance of interrogating vaccine-induced memory B cell responses. The strong correlation of pre-existing antigen-specific memory B cells with post-vaccination serum antibody underscores the immunological connection between memory B cells and recall antibody responses. This relationship likely indicates a role for antigen-specific memory B cells as a source of new antibody secreting cells, as well as potentially contributing to new germinal center responses^18^. Although high circulating titers of neutralizing antibodies are common surrogates of protective immunity, there are many scenarios where circulating antibodies may not achieve sterilizing immunity and additional immune responses will be necessary^19^. For example, high dose viral innoculums may require rapid generation of additional antibody from memory B cells. In addition, if circulating antibodies wane over time, durable memory B cells are likely to provide a rapid source of protective antibody upon antigen re-exposure. Future booster vaccinations, if needed, will focus at least partly on reactivating these antigen-specific memory B cells. Lastly, infection with variant strains that partially escape neutralization by existing circulating antibodies might require strong memory B cell populations that can rapidly re-seed germinal centers and diversify to respond to novel spike antigens^20^. Thus, including analysis of B cell memory in our monitoring of vaccine-induced immune responses not only provides insight into immunological mechanisms of immunity to SARS-CoV2, but may also be useful to inform vaccine implementation decisions across different populations.

## Data Availability

All data is available from the authors upon request.

## ACKNOWLEDGEMENTS

We would like to thank all the subjects who participated in our studies. We also thank the Wherry Lab for helpful discussions.

## CONTRIBUTIONS

RRG, SAA, MMP, DM, ARG, and EJW concieved the study. RRG, SAA, MMP, DM, AP, and SG carried out experiments. RRG, SAA and OK were invovled in clinical recruitment and sample acquisition. All authors participated in data analysis and interpretation. RRG and EJW wrote the manuscript.

## COMPETING INTERESTS

EJW is consulting or is an advisor for Merck, Elstar, Janssen, Related Sciences, Synthekine and Surface Oncology. EJW is a founder of Surface Oncology and Arsenal Biosciences. EJW is an inventor on a patent (US Patent number 10,370,446) submitted by Emory University that covers the use of PD-1 blockade to treat infections and cancer.

**Table 1.**
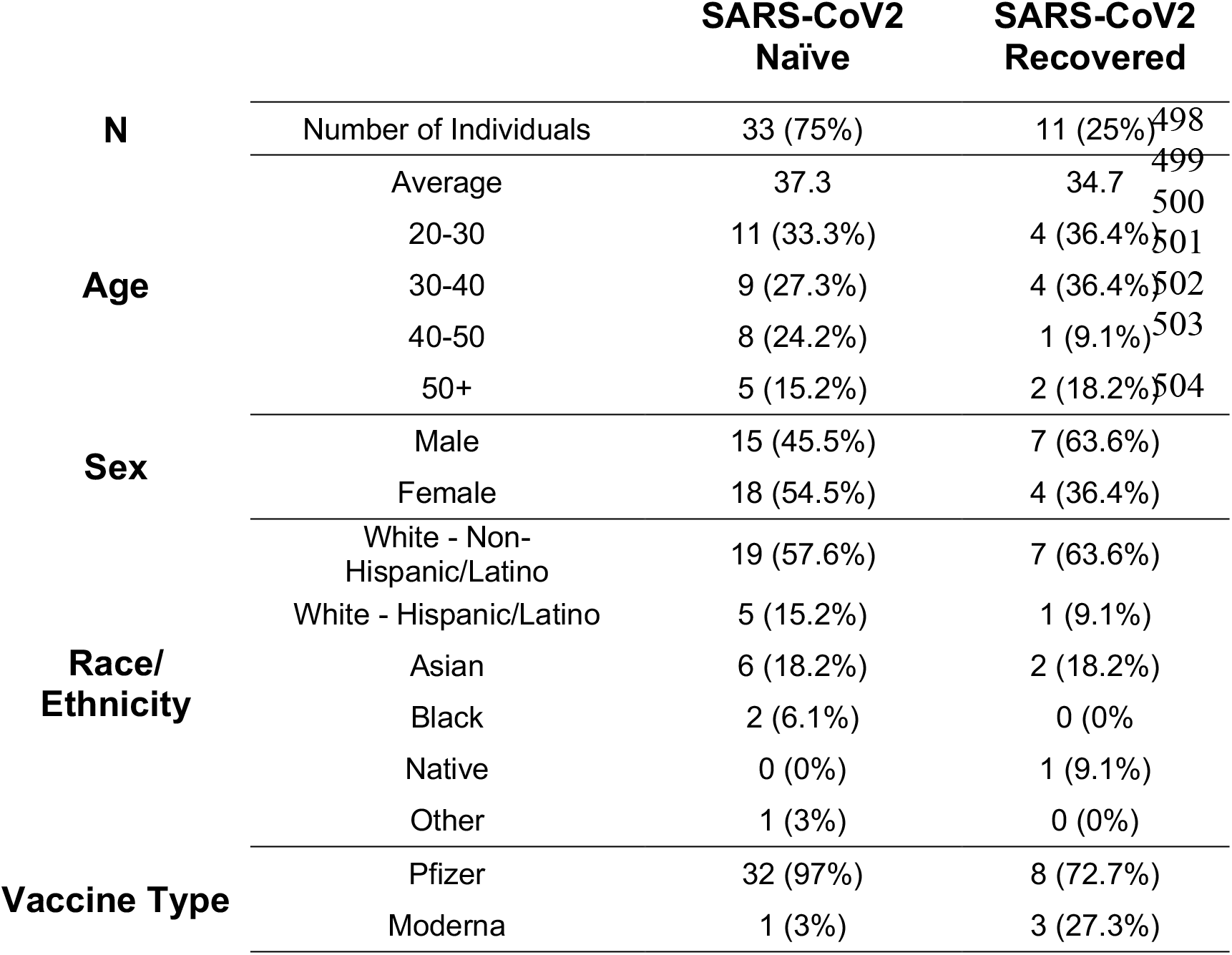
Clinical Characteristics of Individuals Enrolled in the UPenn COVID Vaccine Study.

**Supplemental Figure 1. Gating strategy for antigen-specific B cells**. Lymphocytes were gated by FSC vs. SSC. Doublets were then excluded by FSC-A vs. FSC-H and FSC-A vs. FSC-W. Live cells were identifed as Ghost 510^−^ and total B cells were identified as CD3^-^ CD19^+^. Naïve B cells were then identified as IgD^+^ CD27^-^ and excluded with a boolean not gate. Memory B cells were identified as CD20^+^ CD38^lo/int^ non-naïve B cells. A decoy SA-BV711 probe was used to gate out cells that non-specifically bind streptavidin. Spike-and hemagglutinin-specific B cells were then identified based on their binding to fluorescent probes. Spike^+^ cells were further analyzed for binding to fluorescent RBD probe. Both spike^+^ and spike^+^/RBD^+^ cells were analyzed for IgG vs. IgM expression.

**Supplemental Figure 2. RBD-specificity of spike+ memory B cells**. Frequency of RBD^+^ memory B cells over time in vaccinated individuals. Data are represented as frequency of RBD-specific cells in the spike^+^ memory B cell compartment. RD = non-vaccinated, SARS-CoV2 recovered donors. Dotted lines indicate the mean at baseline in SARS-CoV2 naïve and SARS-CoV2 recovered individuals. Blue = SARS-CoV2 naïve + mRNA vaccine, red = SARS-CoV2 recovered + mRNA vaccine, purple = SARS-CoV2 recovered. Statistics were calculated using unpaired Kruskal-Wallis or Wilcoxon test with adjustment for multiple comparisons.

**Supplemental Figure 3. Sex and age subgroups have increased B cell responses over pre-vaccine baseline. A, B)** Concentration of anti-spike and anti-RBD IgG antibodies at baseline and post-boost compared with sex and age in SARS-CoV2 naïve individuals. Dotted lines indicate the limit of detection for the assay. **C, D)** Frequency of spike^+^ and spike^+^/RBD^+^ memory B cells at baseline and post-boost compared with sex and age in SARS-CoV2 naïve individuals. Data are represented as frequency of spike^+^ and spike^+^/RBD^+^ cells in the total memory B cell compartment. Dotted lines indicate the mean frequency of cells at baseline. **E)** Frequencies of total naïve B, non-naïve B, and memory B cell populations compared with sex and age in SARS-CoV2 naïve individuals.

**Supplemental Figure 4. Association between vaccine-induced side effects and short-term antibody responses. A)** Frequency of self-reported side effects in SARS-CoV2 naïve individuals after the first and second dose of mRNA vaccine. Local side effects include injection site pain, swelling, and redness. Systemic side effects include fever, chills, headache, fatigue, and myalgia. **B)** Concentration of anti-spike and anti-RBD IgG antibodies over time compared with self-reported side effects in SARS-CoV2 naïve individuals. Dotted lines indicate the limit of detection for the assay. **C)** Frequency of spike^+^ and spike^+^/RBD^+^ memory B cells over time compared with self-reported side effects in SARS-CoV2 naïve individuals. Data are represented as frequency of spike^+^ and spike^+^/RBD^+^ cells in the total memory B cell compartment. Post-boost indicates samples collected at timepoint 4 (∼7 days post-secondary vaccination). Statistics were calculated using Wilcoxon test.

