## Supplementary figures and images for "Longitudinal Analysis Reveals Distinct Antibody and Memory B Cell Responses in SARS-CoV2 Naïve and Recovered Individuals Following mRNA Vaccination"

### Supplemental Figures

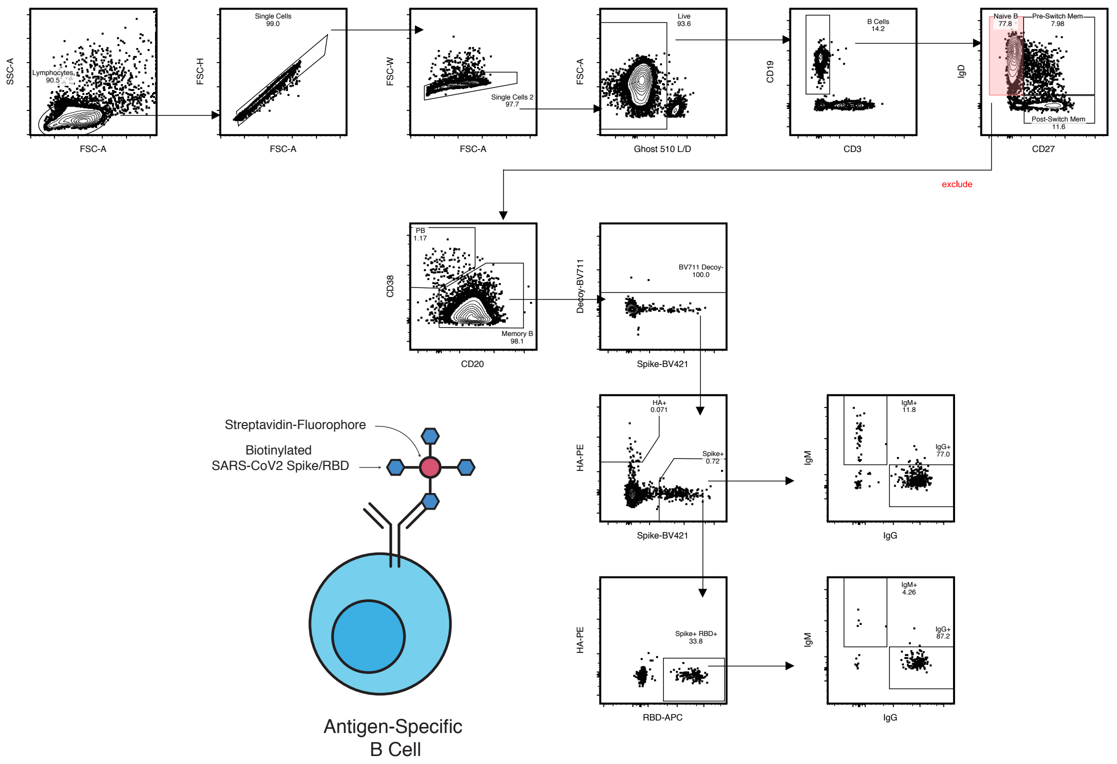

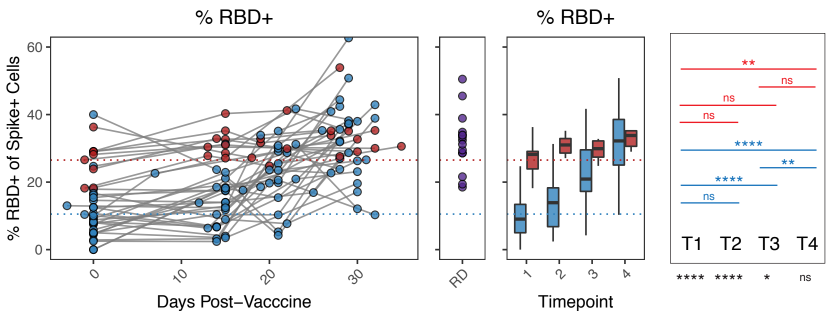

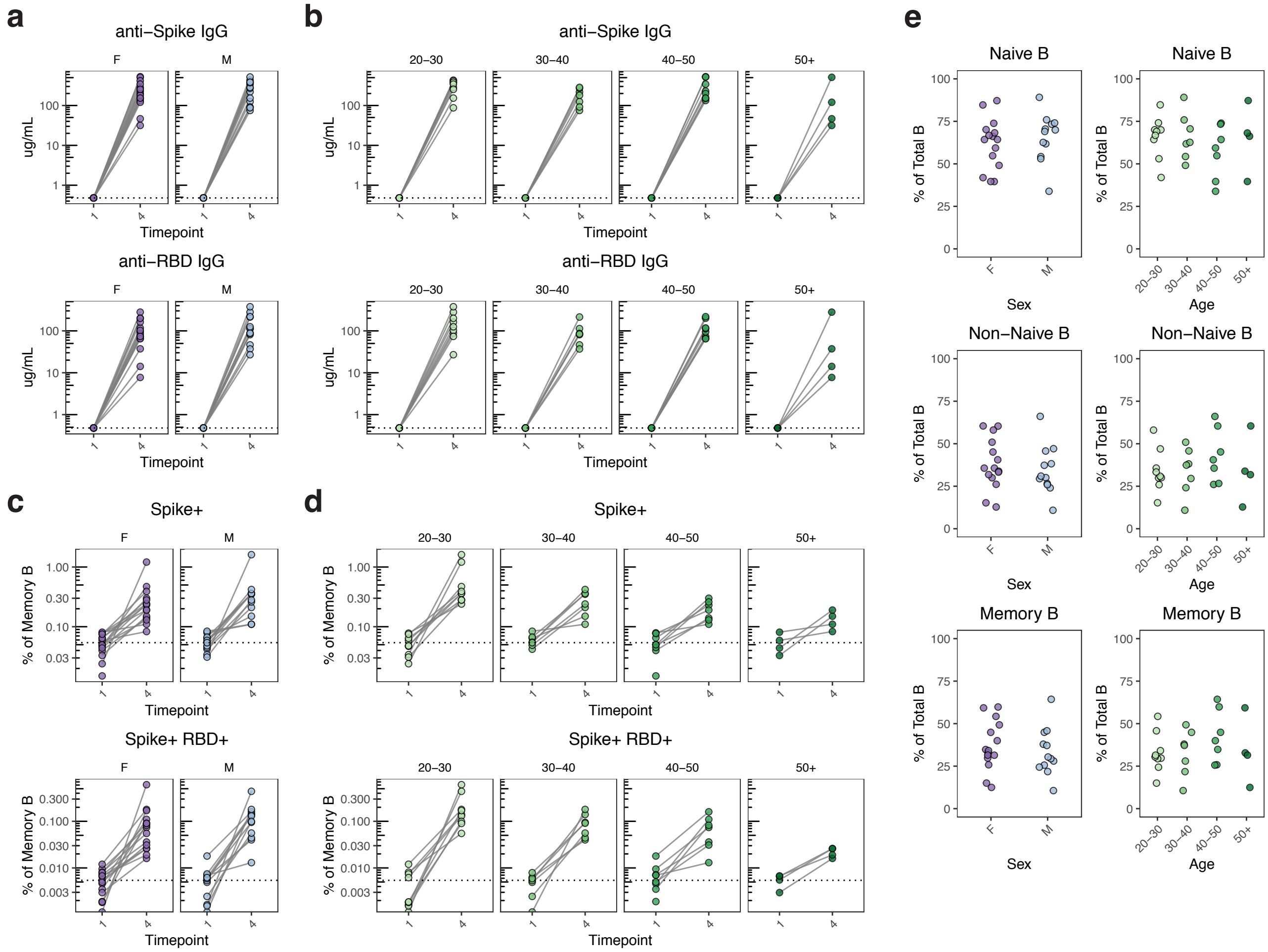

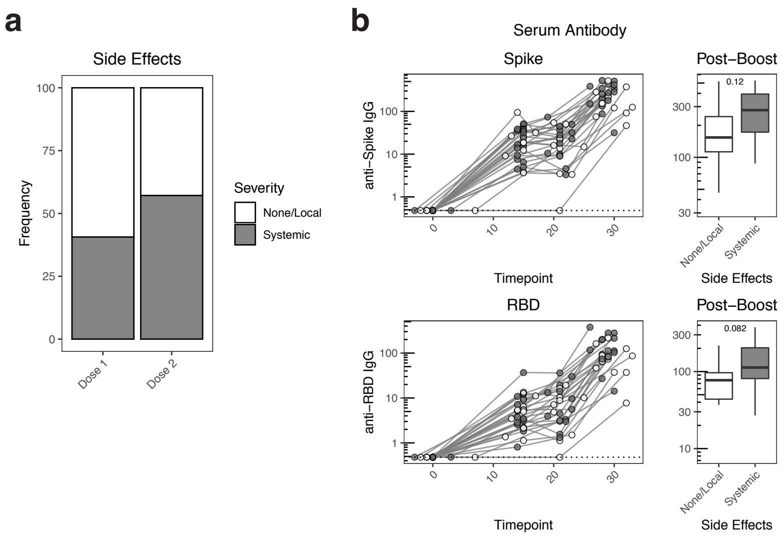
